# Spatiotemporal trends of neglected tropical disease hospitalizations in Ecuador over 25-years from 2000 to 2024

**DOI:** 10.1101/2025.10.28.25338938

**Authors:** Cristina Aldaz-Barreno, Natalia Romero-Sandoval, Monsermín Gualán, Pablo Álvarez, Daniel Zurita-Loma, Gabriela Dávila, Miguel Martin, Philip J Cooper, the Unit on the Social and Environmental Determinants of Health Inequalities (SEDHI)

**Author notes:** Membership of the Unit on the Social and Environmental Determinants of Health Inequalities (SEDHI).

## Abstract

**Background:** World Health Organization has identified 21 neglected tropical diseases (NTDs) affecting millions globally, for control or elimination. We analysed time trends and geographic distribution of hospitalizations attributed to NTDs between 2000 and 2024 in Ecuador.

**Methods:** We estimated hospitalization rates over the 25-year period using hospital discharge data from Ecuadorian national statistics. Time trends Joinpoint regression analysis was done for the 5 most frequent NTDs. Standardized morbidity ratios were estimated for these NTDs for census years (2001, 2010 and 2022) to explore relative changes in hospitalization rates over time and by geography.

**Results:** A total of 179,439 hospital discharges attributed to NTDs were recorded nationally in Ecuador. The five most frequent NTDs accounted for 97.1% of hospitalizations and included arboviral infections, dengue and chikungunya (62% of hospitalizations), snakebite envenoming (20%), soil-transmitted helminthiases (9%), taeniasis and cysticercosis (4%), and scabies and other ectoparasitoses (2%). Only 0.4% of hospitalizations resulted in death. NTD hospitalizations were more frequent in males (54%) with a median age of 19 years (Q_1_ 9 – Q_3_ 37). Although hospitalization rates for all NTDs increased over time, there was a downward trend in non-arboviral NTDs that was significant for snakebite (from 2014, annual percent change -7.81%, 95% CI -11.27, -5.69, P=0.006), soil-transmitted helminths (from 2000, -5.62%, 95% CI -6.56, -4.68, P<0.001), and taeniasis and cysticercosis (from 2003, -10.42%, 95% CI -14.50, -9.68, P=0.002). Relative morbidity caused by NTDs tended to be much greater in Coastal and Amazon provinces, shifting over time particularly to the Amazon region, although taeniasis and cysticercosis morbidity remained greatest in southern Andean Provinces.

**Conclusion:** Hospitalization rates for non-arboviral NTDs have declined over the past 25 years in Ecuador. Populations in the Amazon region remain the most affected by NTD morbidity, highlighting the need for enhanced and appropriately targeted efforts to control and eliminate NTDs.

**Author Summary:** Neglected tropical diseases (NTDs) are a group of diseases affecting millions worldwide, especially in poor and marginalized populations in tropical and subtropical regions of low and middle-income countries. This study analysed hospital records over a 25-year period (2000-2024) in Ecuador—a country where 40% of the population lives in poverty, where there is considerable geoclimatic diversity but where 80% of territory has a tropical or subtropical climate—to determine how hospitalisations due to NTDs have changed over this period and which regions of the country are most affected. Most hospitalisations (97%) were caused by five diseases: the arbovirus infections, dengue and chikungunya; snakebite; intestinal worms; pork tapeworm infections; and scabies. Children and adolescents and those living in the Amazon region suffered most from severe disease caused by NTDs. Over time, hospitalisations from most NTDs declined although those caused by dengue and chikungunya increased. These findings highlight an urgent need to strengthen health education, disease prevention, surveillance, and access to timely diagnosis and treatment tailored to each of these NTDs, especially in the Amazon region of the country. These data help inform the country’s progress meeting its targets for NTD control and elimination by 2030 as outlined in Sustainable Development Goal 3.3.

## Introduction

Neglected Tropical Diseases (NTDs) include a diverse group of 21 conditions caused by parasites, bacteria, viruses, fungi, and non-communicable agents [1]. NTDs were estimated to affect 1.5 billion across 149 countries in 2019 where they caused 150,000 deaths and 19 million DALYs [2]. NTDs are most prevalent in poor and marginalized populations living in tropical and subtropical regions of low and middle-income countries (LMIC) and impose a substantial economic burden on LMIC economies, costing billions of dollars in health costs annually [3,4].

Optimal interventions against NTDs diseases are addressed under Target 3.3 of the United Nations Sustainable Development Goals (SDGs). WHO’s *NTD Roadmap 2021– 2030* sets a target of reducing by 90% the global population requiring interventions for NTDs by 2030 [1]. Using 2021 as the baseline year, the WHO outlined several strategic goals for NTDs by 2030: control measures for nine, elimination as a public health problem for six, interruption of transmission for three, and complete eradication for the remaining three [5]. An estimated 51 million people in Latin America required NTDs-related interventions in 2022 [6].

Hospitalizations due to NTDs represent the most severe end of the clinical spectrum for these diseases including acute complications or comorbidities indirectly linked to NTDs [7–10]. High rates of NTD hospitalizations place a significant economic strain on impoverished families and communities, further exacerbating preexisting vulnerabilities relating to poverty, limited access to healthcare, and discrimination based on ethnicity, language, or culture [11].

Ecuador, an upper-middle-income country of 17 million on the Pacific coast of South America, currently monitors seven notifiable NTDs [12] including rabies, dengue and chikungunya, Chagas disease, snakebite envenoming, leishmaniasis, leprosy, and onchocerciasis. Significant progress has been made in Ecuador to meet SDG 3.3 for some NTDs - the formal certification of the interruption of transmission of onchocerciasis was completed in 2015 [13] and is pending for yaws [14,15] - while for others, there are more limited published data at a national level on current status [16], or temporal trends or geographic distribution within the country (myiasis [17]), dengue [18–21], cysticercosis [22], and Chagas [19,20,23]. Most published data on NTDs have been limited to studies in geographically restricted populations in areas known to be highly endemic for these diseases [15,17,23,24].

The objective of the present study was to describe time trends nationally and standardised rates by province of hospitalizations attributed to NTDs in Ecuador over a 25-year period between 2000 and 2024. The goal was to improve our understanding of the spatiotemporal epidemiology of severe morbidity attributed to NTDs (i.e., requiring hospitalization) to inform appropriate public health interventions for their control and elimination and contribute to assessing the country’s progress in meeting SDG target 3.3 relating to NTDs.

## Methods

### Study Design

We conducted an ecological study to analyse time trends and spatial patterns of hospitalizations due to NTDs in Ecuador from 2000 to 2024.

### Setting and selection of ICD codes

Ecuador is an upper-middle income country with high levels of inequality and where approximately 40% of the population are classified as living in poverty [25]. The country has a population of approximately 17 million and has considerable geoclimatic diversity. Ecuador lies on the equator and is bisected north-south by the Andes, dividing the country into 4 distinct geoclimatic regions (Coast, Andes, Amazon, and the Galápagos archipelago [Insular]). The country experiences wet and dry seasons that vary by month and intensity between geoclimatic regions. Tropical and subtropical areas, that account for approximately 87% of the country’s land area [26], provide favourable conditions for the survival of disease vectors and the transmission of infectious pathogens.

Ecuador is divided administratively into 24 provinces, grouped into four distinct geoclimatic regions: six in the Coastal region, ten in the Andean region, five in the Amazon region, and one representing the Galápagos archipelago. The national health system operates under an integrated network model based around primary health care. All inpatient healthcare facilities—public and private —are required to report hospital discharges to the National Institute of Statistics and Censuses (INEC) using ICD-10 codes. This is done via a standardized process based on the primary diagnosis recorded by the healthcare provider. INEC is responsible for data quality control, storage, and dissemination of data for administrative and research use [27].

### Variables and source of hospitalization data

The 21 NTDs defined by WHO are: Buruli ulcer; Chagas disease; dengue and chikungunya (dengue/chik); dracunculiasis; echinococcosis; foodborne trematodiases; human African trypanosomiasis; leishmaniasis; leprosy; lymphatic filariasis; mycetoma, chromoblastomycosis and other deep mycoses; noma; onchocerciasis; rabies; scabies and other ectoparasitoses; schistosomiasis; soil-transmitted helminthiases; snakebite envenoming; taeniasis/cysticercosis; trachoma; and yaws [28]. For this analysis, NTDs were divided into two groups: i) the first included 14 NTDs (referred here to as endemic NTDs) considered endemic or with a known historical presence in Ecuador and included: dengue/chik; snakebite envenoming (snakebite); soil-transmitted helminths (STH); taeniasis and cysticercosis (taeniasis/cysticercosis); scabies and other ectoparasitoses (ectoparasitoses); and less frequent NTDs (named, Other NTDs)— including Chagas disease, echinococcosis, leishmaniasis, leprosy, onchocerciasis, yaws, foodborne trematodiases, mycetoma, chromoblastomycosis and other deep mycoses, and rabies; and ii) the second group (referred to as non-endemic NTDs) were those considered not to be or have been endemic in the country and included Buruli ulcer, drancunculiasis, human African trypanosomiasis, lymphatic filariasis, noma, schistosomiasis, and trachoma. ICD-10 codes corresponding to NTDs (S1 Table) were used to identify relevant hospital discharge diagnoses from 2000 to 2024 using the national hospital discharge registry, that covers the period 2000 to 2024 [29] and which provides aggregated monthly and annual data at provincial level. This registry provides also nominal data on sex, health status at discharge (alive or dead), age, duration of hospitalization, ethnicity (from 2014), and area of usual residence (from 2015). Dengue was classified under codes A90–A92 for the entire study period, with A97 added in 2019 following WHO recommendations [30]. Endemic NTDs were analysed at national and provincial levels. For non-endemic NTDs, area of usual residence was categorised into 3 relevant geographic areas: Ecuador, South America (outside Ecuador), and Africa.

### Statistical analysis

Descriptive statistics (absolute and relative frequencies) were calculated for the endemic NTDs (dengue/chik, snakebite, STH, taeniasis/cysticercosis, ectoparasitoses, and Other NTDs) and stratified by demographic variables. Age-standardized rates (ASR) and 95% confidence intervals (95% CI) were calculated per 100,000 population for each endemic NTD by the direct method based using the WHO world standard population [31]. Trends in NTD hospitalizations over the study period were assessed using Joinpoint regression models [32]. This method identifies significant shifts in trend by linking multiple linear segments on a logarithmic scale at specific points known as “joinpoints.” The analysis begins with a model assuming no joinpoints (i.e., a single straight line) and incrementally adds points where statistically significant changes in the slope are detected over time. The Joinpoint technique calculates the Annual Percentage Change (APC) between these inflection points and estimates the Average Annual Percentage Change (AAPC) across the entire study duration. Statistical testing determines whether each APC significantly deviates from zero. In the final model, each joinpoint signifies a significant alteration in the direction or magnitude of the trend, with corresponding APC values. If no joinpoints are found, the APC remains constant and is equivalent to the AAPC. APC for the five most prevalent NTDs was estimated and Joinpoint regression used to detect significant trends. A seasonality index for dengue/chik and snakebite was calculated by aggregating monthly hospitalizations between 2000 to 2024 and assigning a value of 1 where frequency was equal to the annual average, >1 where frequency was above the annual average, and <1 when below the annual average as described [33,34]. We estimated crude hospitalization rates stratified by sex and age group for each province for 2000 and 2024 using population projections [35]. To compare hospitalization rates across provinces, we calculated standardized morbidity ratios (SMRs) and their 95% confidence intervals for the 3 census years (2000, 2010, 2022) using indirect standardization with the national census population as reference [36]. SMR maps were created in QGIS 1.38 using fixed interval scales across NTDs and census years. Non-endemic NTDs were summarized using absolute frequencies per year and by origin (i.e. Ecuador, outside Ecuador but in South America, and Africa) from 2015 when data became available. Analyses were done using SPSS (V29.0.1.0), R Studio, Joinpoint Regression Program (V5.0.2), and QGIS software.

### Ethical Considerations

This study used anonymized secondary publicly accessible data.

## Results

Between 2000 and 2024 a total of 179,439 hospitalizations, representing 0.7% of all hospitalizations, were attributed to NTDs through statutory reporting to the INEC. Table 1 summarizes national frequencies of hospital discharges for endemic NTDs, stratified by key sociodemographic variables. These include ethnicity (from 2014), length of hospital stay, health status at discharge, and urban–rural residence (from 2015). Arboviral infections (dengue/chik) were the most common, representing 62.4% of NTD hospitalizations, followed by snakebite (20.1%), STH (8.7%), taeniasis/cysticercosis (3.9%), and ectoparasitoses (2.0%). The other nine non-endemic NTDs each contributed <1%, together accounting for just 2.1% of cases.

**Table 1.** Sociodemographic characteristics and hospital stay characteristics of hospital discharges of all 21 neglected tropical diseases (NTDs) and the 14 endemic NTDs in Ecuador between 2000 and 2024.

|  | Total (%) | Sex |  | Age (years) | Health status at discharge |  | Hospital stay (days) | Area of residence* (n=74,880) |  | Minorities*<br>* (n=84,849) |
| --- | --- | --- | --- | --- | --- | --- | --- | --- | --- | --- |
|  |  | Men<br>n (%) | Women<br>n (%) | Median (Q1-Q3) | Alive<br>n (%) | Dead<br>n (%) | Median (Q1-Q3) | Urban<br>n (%) | Rural<br>n (%) | n (%) |
| All NTDs | 179439 (100) | 96664 (53.9) | 82775 (46.1) | 19 (9-37) | 178632 (99.6) | 807 (0.4) | 3 (2-5) | 61930 (82.7) | 12950 (17.3) | 7231 (8.5) |
| Endemic NTDs |  |  |  |  |  |  |  |  |  |  |
| Dengue and chikungunya | 111946 (62.4) | 57126 (51) | 54820 (49) | 16 (9-31) | 111570 (99.7) | 376 (0.3) | 3 (2-5) | 47167 (88.3) | 6262 (11.7) | 2632 (4.4) |
| Snakebite envenoming | 36070 (20.1) | 23354 (64.7) | 12716 (35.3) | 28 (15-46) | 35906 (99.5) | 164 (0.5) | 3 (2-5) | 7515 (61) | 4807 (39) | 3332 (23.4) |
| Soil-transmitted helminthiases | 15647 (8.7) | 7450 (47.6) | 8197 (52.4) | 10 (4-28) | 15619 (99.8) | 28 (0.2) | 3 (2-4) | 3445 (75.9) | 1096 (24.1) | 975 (18.9) |
| Taeniasis and cysticercosis | 6986 (3.9) | 3595 (51.5) | 3391 (48.5) | 42 (28-58) | 6906 (98.9) | 80 (1.1) | 4 (3-8) | 1084 (82.7) | 226 (17.3) | 56 (3.7) |
| Scabies and other ectoparasitoses | 3569 (2) | 2027 (56.8) | 1542 (43.2) | 13 (4-57) | 3526 (98.8) | 43 (1.2) | 5 (2-8) | 1471 (83.6) | 289 (16.4) | 122 (6.3) |
| Leishmaniasis | 1145 (0.6) | 852 (74.4) | 293 (25.6) | 26 (19-48) | 1142 (99.7) | 3 (0.3) | 4 (1-9) | 253 (71.5) | 101 (28.5) | 54 (13.9) |
| Leprosy | 1019 (0.6) | 718 (70.5) | 301 (29.5) | 55 (43-68) | 980 (96.2) | 39 (3.8) | 25 (9-92) | 42 (77.8) | 12 (22.2) | 5 (6.5) |
| Mycoses | 628 (0.3) | 344 (54.8) | 284 (45.2) | 35 (16-52) | 612 (97.5) | 16 (2.5) | 2 (1-9) | 321 (87.7) | 45 (12.3) | 16 (3.7) |
| Chagas disease | 494 (0.3) | 309 (62.6) | 185 (37.4) | 62.5 (38-77) | 461 (93.3) | 33 (6.7) | 5 (2-10) | 144 (85.2) | 25 (14.8) | 16 (8.2) |
| Echinococcosis | 253 (0.1) | 101 (39.9) | 152 (60.1) | 40 (23-60) | 251 (99.2) | 2 (0.8) | 5 (3-11) | 108 (76.1) | 34 (23.9) | 6 (3.9) |
| Yaws | 230 (0.1) | 111 (48.3) | 119 (51.7) | 31 (15-52) | 229 (99.6) | 1 (0.4) | 1 (1-3) | 104 (90.4) | 11 (9.6) | 5 (3.8) |
| Foodborne trematodiasis | 154 (0.1) | 76 (49.4) | 78 (50.6) | 25 (12-41) | 152 (98.7) | 2 (1.3) | 5 (2-8) | 28 (77.8) | 8 (22.2) | 6 (14.3) |
| Rabies | 83 (0) | 19 (22.9) | 64 (77.1) | 26 (20-35) | 76 (91.6) | 7 (8.4) | 2 (1-3) | 2 (100) | 0 (0) | 0 (0) |
| Onchocerciasis | 8 (0) | 2 (25) | 6 (75) | 19 (7-55.5) | 8 (100) | 0 (0) | 2 (1.5-6) | 1 (100) | 0 (0) | 0 (0) |

\* Data available from 2015
\*\* Data available from 2014
All NTDs – any of the 21 NTDs recognised by WHO.
Minorities – proportion belonging to any of the ethnic groups: Indigenous, Afro-Ecuadorian, and Montubio.
Mycoses – Mycetoma, chromoblastomycosis and other deep mycoses.

NTD hospitalizations tended to be more common in males (53.9%), particularly for snakebite (64.7% of hospitalizations). Mortality was low – only 0.4% of NTD hospitalizations resulted in the death of the patient although this was much higher for Chagas disease (6.7%). NTD hospitalizations were much more frequent for populations whose area of habitual residence was urban (82.7% of hospitalizations) and was affected by the strong urban bias for arbovirus infections (88.3% of hospitalizations). Only 8.5% of hospitalizations were reported for ethnic minorities (i.e., Indigenous, Montubio, and Afro-Ecuadorian), much lower than the population proportion in the 2022 national census (20.2%) [25].

Cumulative annual age-standardized rates for endemic NTDs in Ecuador are shown in Figure 1 (see S2 Table for annual rates with 95% confidence intervals and S1 Fig for annual rates for specific NTDS). Hospitalization rates varied markedly over time, largely driven by fluctuations in dengue/chik. Table 2 shows joinpoint trends in age-standardized hospitalization rates per 100,000 population for the 5 most frequent endemic NTDs in Ecuador between 2000 and 2024. The average annual percentage change (AAPC) for all NTDs was 2.66% (95% CI 0.75-5.03) and for dengue/chik 7.79% (95% CI 4.23-12.32), with the other NTDs showing a downward trend.

**Fig 1.**
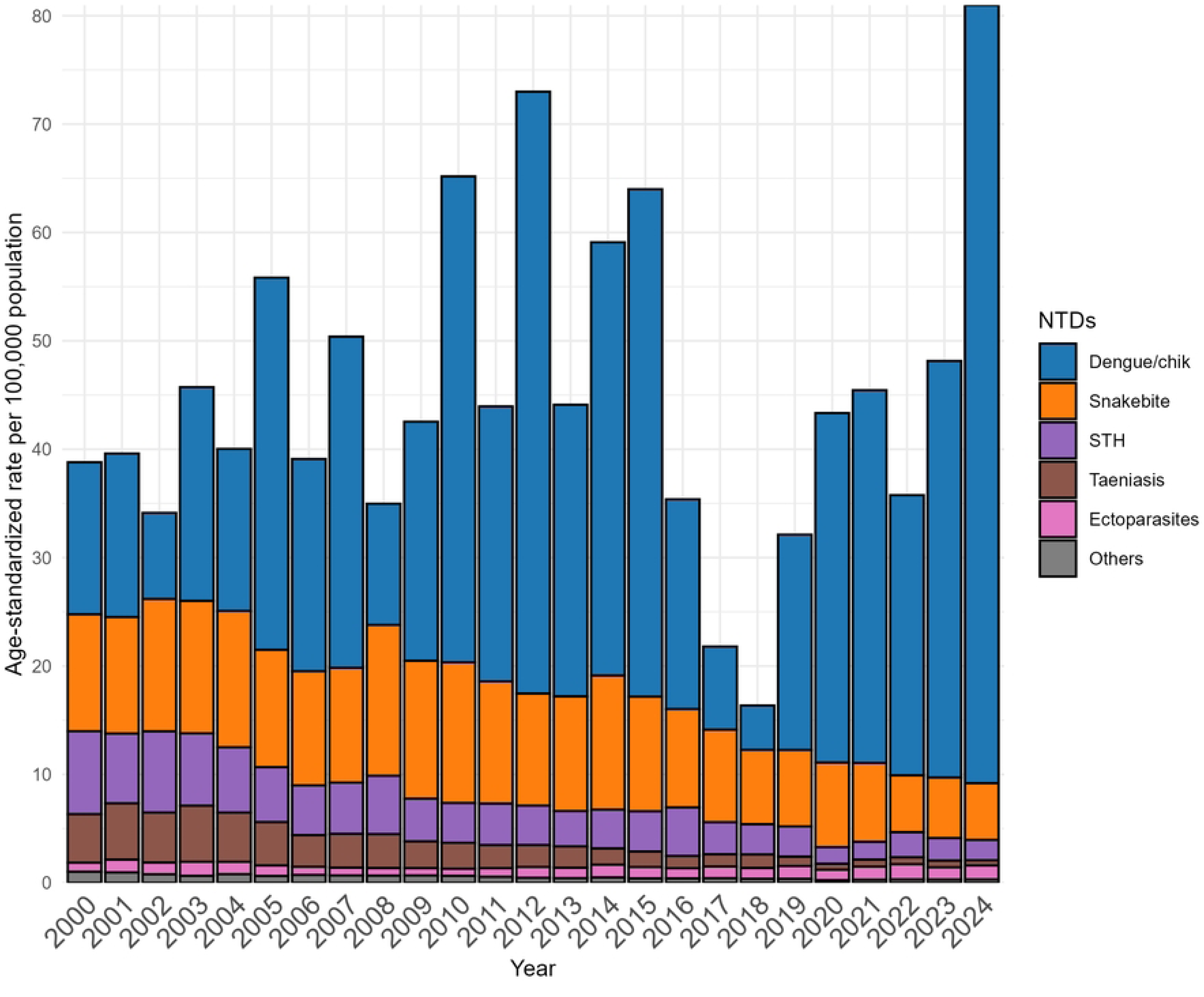
Cumulative annual age-standardized hospitalizations rates (per 100,000 population) attributed to endemic NTDs in Ecuador between 2000 and 2024. The contribution of the 14 NTDs grouped into 6 disease groups: Dengue and chikungunya, Snakebite envenoming, Soil-transmitted helminths, Taeniasis and cysticercosis, Scabies and other ectoparasitoses, and Other NTDs — including Chagas disease, echinococcosis, leishmaniasis, leprosy, onchocerciasis, yaws, foodborne trematodiases, mycetoma, chromoblastomycosis and other deep mycoses, and rabies.

**Table 2.** Joinpoint trends in age-standardized hospitalization rates (per 100,000 population) for all 21 neglected tropical diseases (NTDs) and the 14 endemic NTDs in Ecuador between 2000 and 2024.

| NTD | Period, 2000-2024 |  | Trend 1 |  |  | Trend 2 |  |  | Trend 3 |  |  |
| --- | --- | --- | --- | --- | --- | --- | --- | --- | --- | --- | --- |
|  | AAPC<br>(95% CI) | P<br>value | Years | APC<br>(95% CI) | P<br>value | Years | APC<br>(95% CI) | P<br>value | Years | APC<br>(95% CI) | P<br>value |
| All 21 NTDs | 2.66<br>(0.74-5.03) | 0.018 | 2000-2015 | 2.72<br>(0.22-7.86) | 0.037 | 2015-2018 | -29.60<br>(-39.13—7.22) | 0.007 | 2018-2024 | 23.78<br>(11.61-75.51) | 0.008 |
| Dengue and<br>chikungunya | 7.76<br>(4.23 – 12.32) | 0.002 | 2000-2015 | 8.16<br>(3.22 – 18.97) | 0.008 | 2015-2018 | -43.75<br>(-56.48 –8.13) | 0.009 | 2018-2024 | 47.78<br>(22.36 – 160.11) | 0.006 |
| Snakebite<br>envenoming | -3.45<br>(-4.34 - -2.62) | <0.001 | 2000-2014 | -0.21<br>(-1.64 – 1.93) | 0.835 | 2014-2024 | -7.81<br>(-11.27 - -5.69) | <0.001 |  |  |  |
| Soil-transmitted<br>helminths | -5.62<br>(-6.56 –4.68) | <0.001 | 2000-2024 | -5.62<br>(-6.56 –4.68) | <0.001 |  |  |  |  |  |  |
| Taeniasis and<br>cysticercosis | -8.89<br>(-10.47 - -7.57) | <0.001 | 2000-2003 | 2.62<br>(-9.21 – 27.96) | 0.732 | 2003-2024 | -10.42<br>(-14.50 - -9.68) | 0.002 |  |  |  |
| Scabies and<br>ectoparasitose<br>s | 1.69<br>(0.62 – 3.18) | 0.023 | 2000-2003 | 13.63<br>(-3.33 – 39.11) | 0.072 | 2003-2007 | -14.53<br>(-23.28 – 4.13) | 0.065 | 2007-2024 | 3.88<br>(2.29 – 5.73) | 0.022 |

The Annual Percent Change (APC) and Average Annual Percentage Change (AAPC) is significantly different from zero (P<0.05).
CI – confidence interval.

Trends in hospitalization rates for all NTDs and for the five most common endemic NTDs with joinpoint regression lines are shown in Figure 2. Trends in hospitalization rates were strongly affected by arboviral hospitalization rates (Fig 2A and B) with an increase observed over the period 2000-2015 (dengue/ckik, APC 8.16, 95% CI 3.22-19.97), followed by a decline between 2015 and 2018 (dengue/chik, APC -43.75, 95% CI -56.48 - -8.13), and a large increase between 2018 and 2024 (dengue/chik, APC 47.78, 95% CI 22.36-160.11). Snakebite envenoming showed a significant decrease between 2014 and 2024 (APC -7.81, 95% CI -11.27- -5.69) (Fig 2C), soil-transmitted helminths declined over the whole observation period (AAPC -5,62, 95% CI -6.56 - - 4.68) (Fig 2D), while taeniasis and cysticercosis declined significantly between 2003 and 2024 (APC, -10.42, 95% CI -14.5 - -9.68) (Fig 2E). Scabies and other ectoparasitoses appeared to show a significant increase from 2007 (APC, 3.88, 95% CI 2.29-5.73) (Fig 2F). Annual reported number of hospitalizations attributed to non-endemic NTDs are shown in S3 Table with area of usual residence for patients with these NTDs provided in S4 Table.

**Fig 2.**
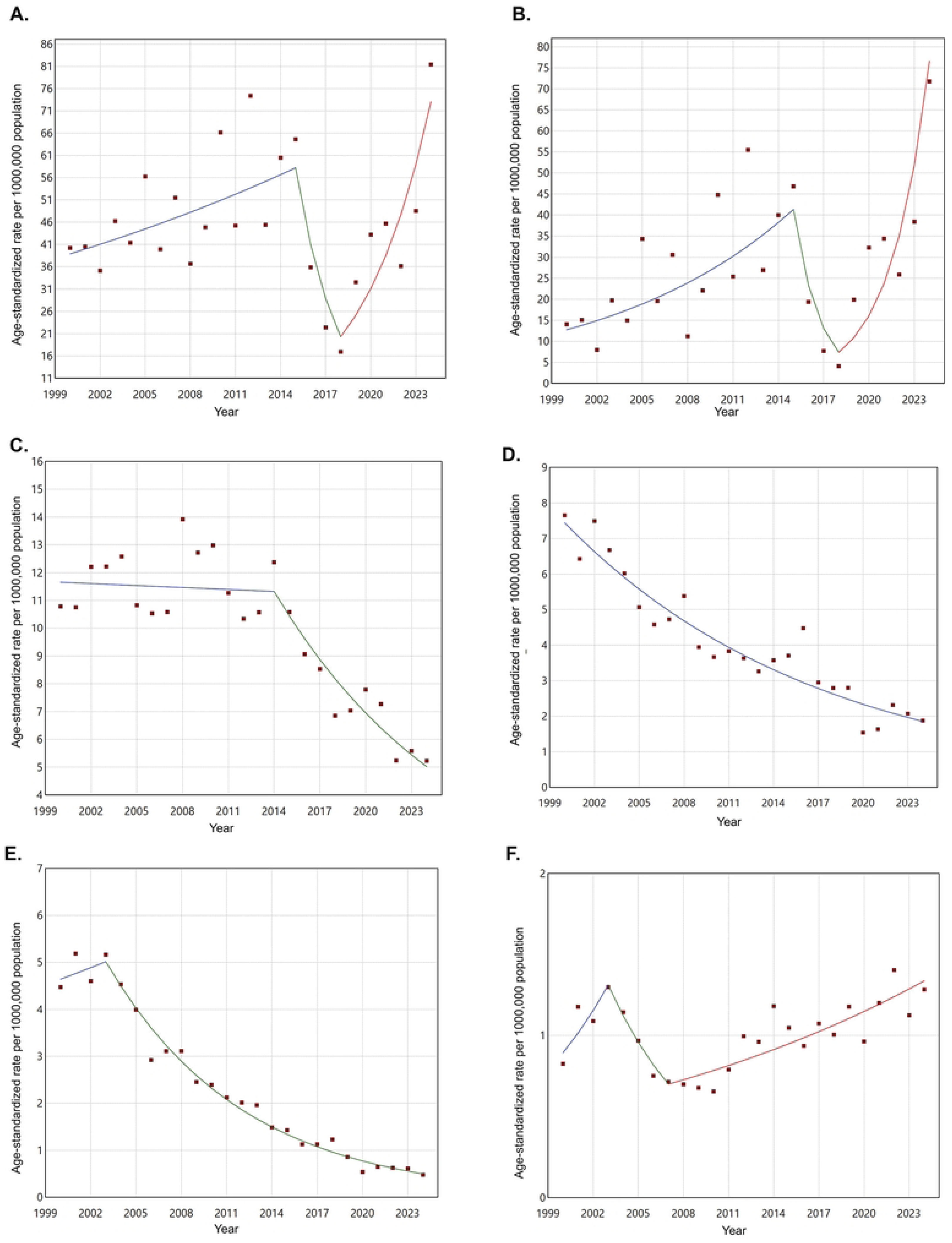
Trends of age-standardized hospitalizations rates (per 100,000 population) attributed to NTDs in Ecuador between 2000 and 2024. Shown are hospitalization rates NTDs and for the 5 most frequent endemic NTDs with joinpoint regression lines. Shown are hospitalization rates NTDs and for the 5 most frequent endemic NTDs in Ecuador with joinpoint regression lines. (A) All NTDs, 21 Neglected Tropical Diseases; (B) Dengue and chikungunya; (C) Snakebite envenoming; (D) Soil-transmitted helminths; (E) Taeniasis and cysticercosis; (F) Scabies and other ectoparasitoses.

Figure 3 shows how hospitalization frequencies for dengue/chik (Fig 3A) and snakebite (Fig 3B) varied monthly, reported at five-year intervals between 2000 and 2024, as well as the seasonality indices for these hospitalizations (Fig 3C and 3D, respectively). Clear seasonality (index >1) was observed for dengue/chik between March and July and for snakebite between February and July. The other endemic NTDs showed no clear evidence of seasonality.

**Fig 3.**
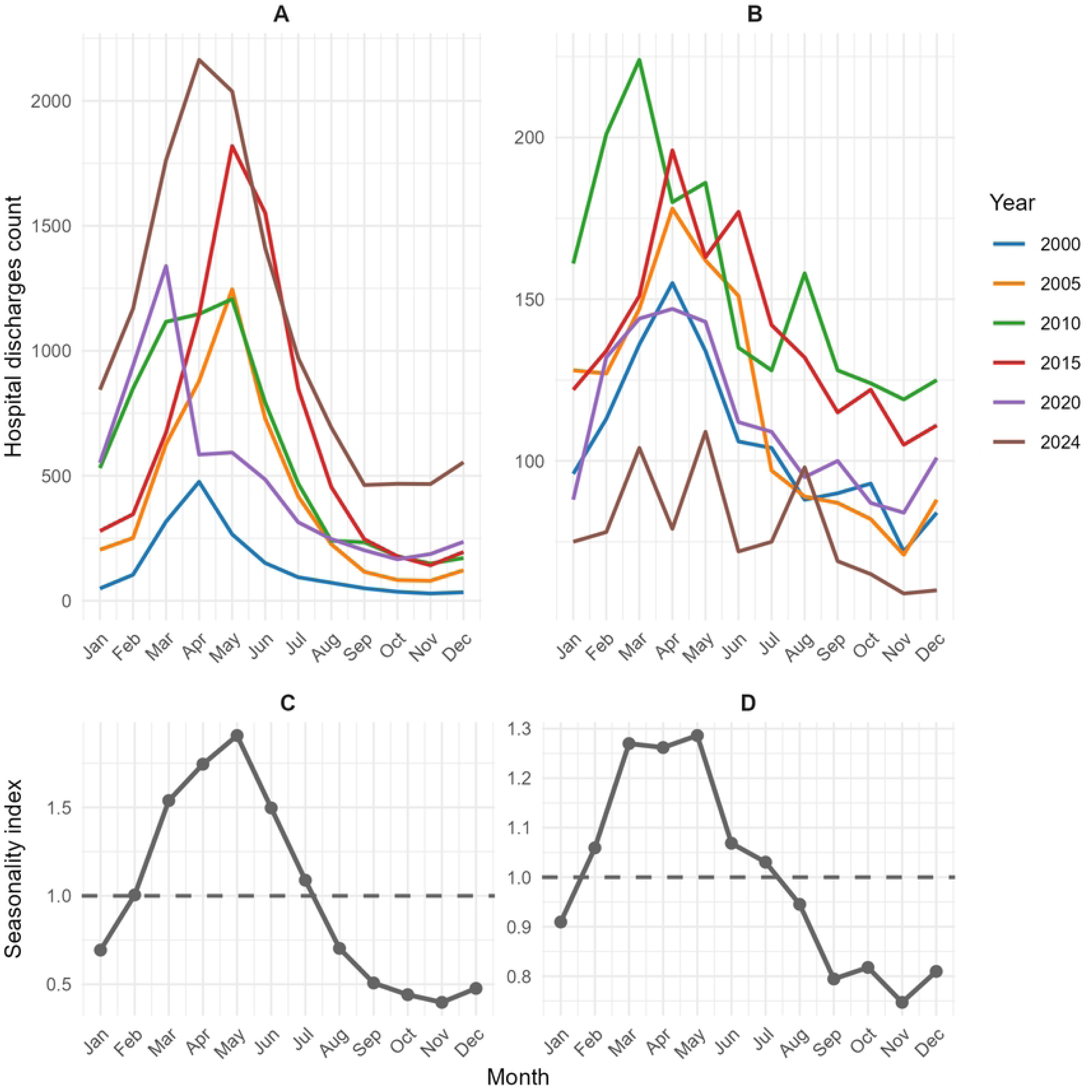
Monthly frequencies and seasonality indices for hospitalizations attributed to the arboviral infections, dengue and chikungunya (A & C), and snakebite envenoming (B & D) in Ecuador between 2000 and 2024. Lines represent frequencies reported at five-year intervals according to colour code shown in key.

Figures 4A and 4B shows crude hospitalization rates for 14 endemic NTDs by sex and age group in 2000 and 2024, while Figures 5A and 5B show these distributions for non-arboviral NTDs. Greater rates of endemic NTDs were observed in 2024, largely among those aged below 30 years, but these greater rates were attributed to the arboviral infections. Non-arboviral NTD hospitalization rates tended to increase with age and were reduced across all ages in 2024 compared to 2000, and rates appeared to be greater in men. Scabies and other ectoparasitoses had emerged by 2024 as an important cause of NTD hospitalizations, particularly among the elderly.

**Fig 4.**
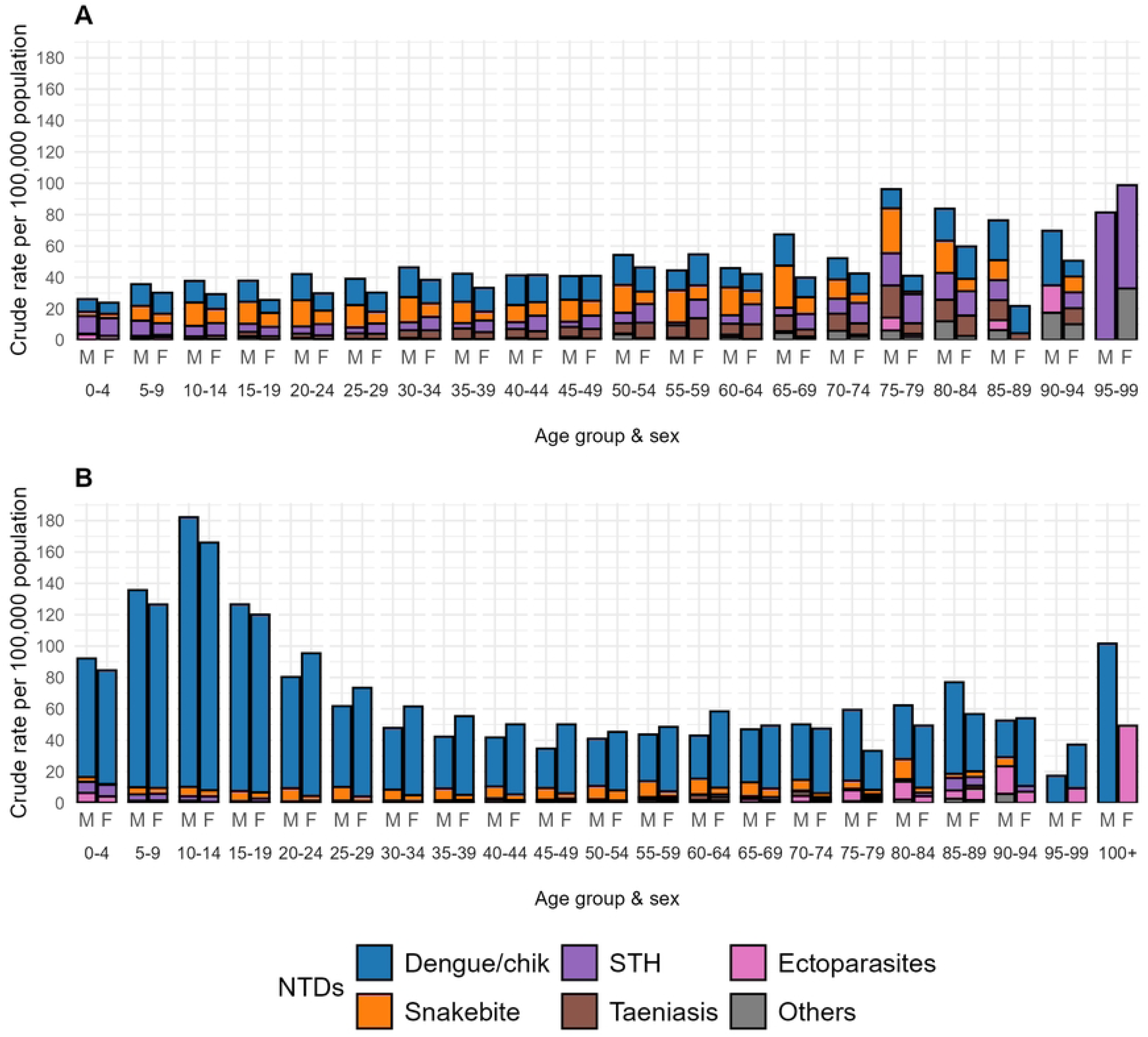
Crude cumulative hospitalization rates (per 100,000 population) stratified by sex and age grouping for NTDs in Ecuador during the years 2000 (A) and 2024 (B). Shown are data for the 5 most frequent NTDs and all others. M = male, F = female; **(A)** Crude rates for 14 endemic NTDs grouped into 6 disease groups: Dengue and chikungunya; Snakebite envenoming; Soil-transmitted helminths; Taeniasis and cysticercosis; Scabies and other ectoparasitoses; and Other NTDs — including Chagas disease, echinococcosis, leishmaniasis, leprosy, onchocerciasis, yaws, foodborne trematodiases, mycetoma, chromoblastomycosis and other deep mycoses, and rabies from 2001; **(B)** All endemic NTDs from 2024.

**Fig 5.**
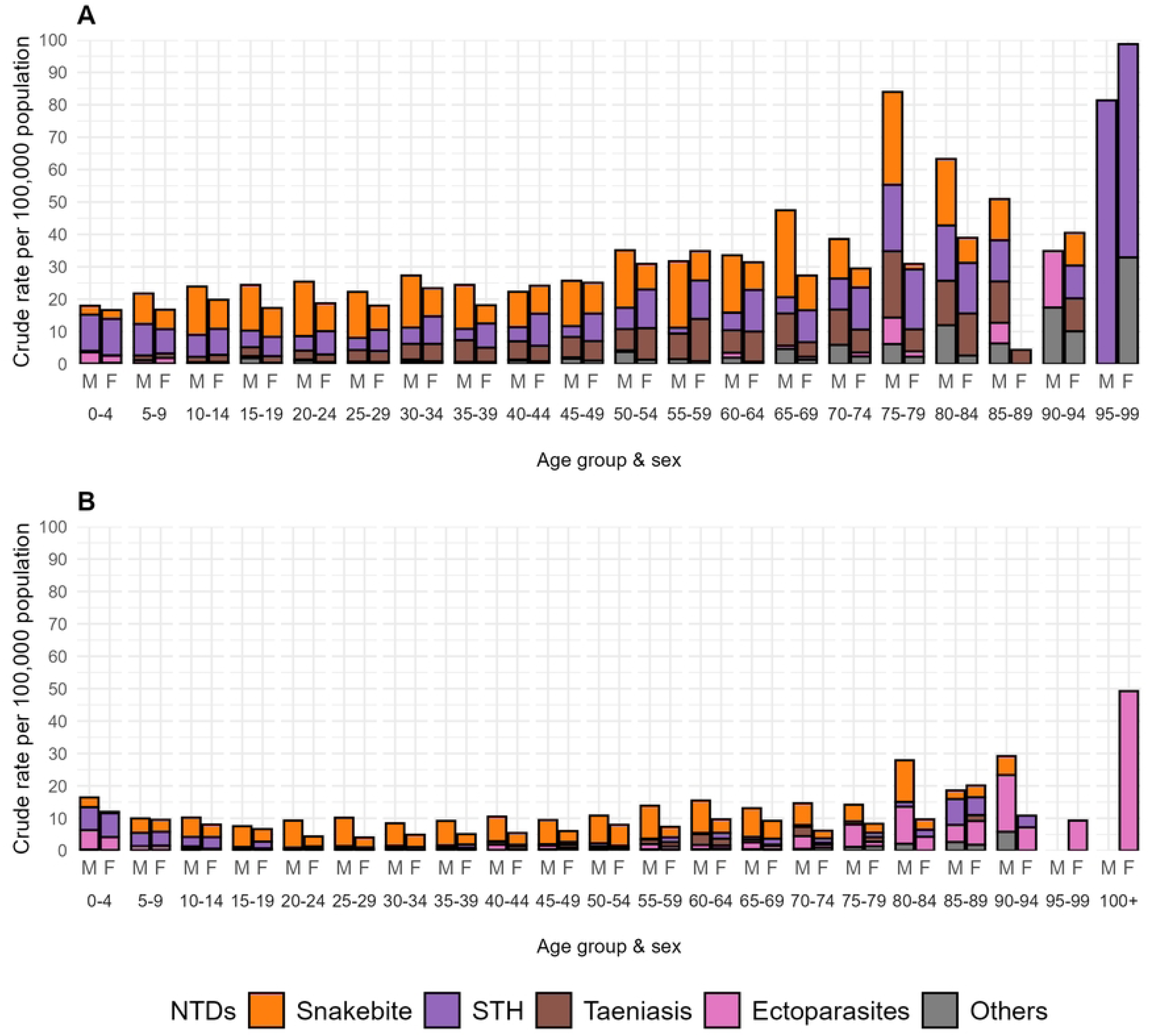
Crude cumulative hospitalization rates (per 100,000 population) stratified by sex and age grouping for non-arboviral NTDs for 2000 (A) and 2024 (B). Shown for the 4 most frequent non-arboviral NTDs and all others (non-arboviral). M = male, F = female; A and B show crude rates for all non-arboviral endemic NTDs in 2000 and 2024, (1) Snakebite envenoming; (2) Soil-transmitted helminths; (3) Taeniasis and cysticercosis; (4) Scabies and other ectoparasitoses; (5) and **Other NTDs** — including Chagas disease, echinococcosis, leishmaniasis, leprosy, onchocerciasis, yaws, foodborne trematodiases, mycetoma, chromoblastomycosis and other deep mycoses, and rabies.

Data for the hospitalizations caused by endemic NTDs by Province in 2000 and 2024 are shown in S2 Fig. In 2000, rates were highest in the Amazon provinces (Morona Santiago Napo, Orellana, Pastaza, and Zamora Chinchipe) with particularly high rates of snakebite. A similar pattern was observed in 2024, except for the province of Napo, which had the highest rate in two years.

We estimated standardized morbidity ratios for the 5 most frequent endemic NTDs for each of the 3 census years by province (2001, 2010, 2022). The data are provided in S5 Table and illustrated graphically in Figure 6. Relative morbidity was shown as white (expected level), shades of increasing blue (increasingly lower than expected), and shades of increasing red (increasingly greater than expected). Dengue/chik (Figure 6A) hospitalizations were lower than expected in the Andean provinces over the 3 census years and tended to be greater than expected in Coastal provinces and over time in the Amazon provinces. For snakebite (Figure 6B) and STH (Figure 6C), the Amazon region had a much higher than expected risk while the Andean provinces had a lower risk over the observation period. In the case of taeniasis/cysticercosis (Fig 6D), the risk was consistently higher in Andean, particularly southern Andean provinces. Ectoparasitoses (Fig 6E) showed less clear trends with excess morbidity risk shifting from central and northern Amazon provinces in 2001 to southern Amazon/Andean provinces in 2010, to central and northern coastal provinces in 2022.

**Fig 6.**
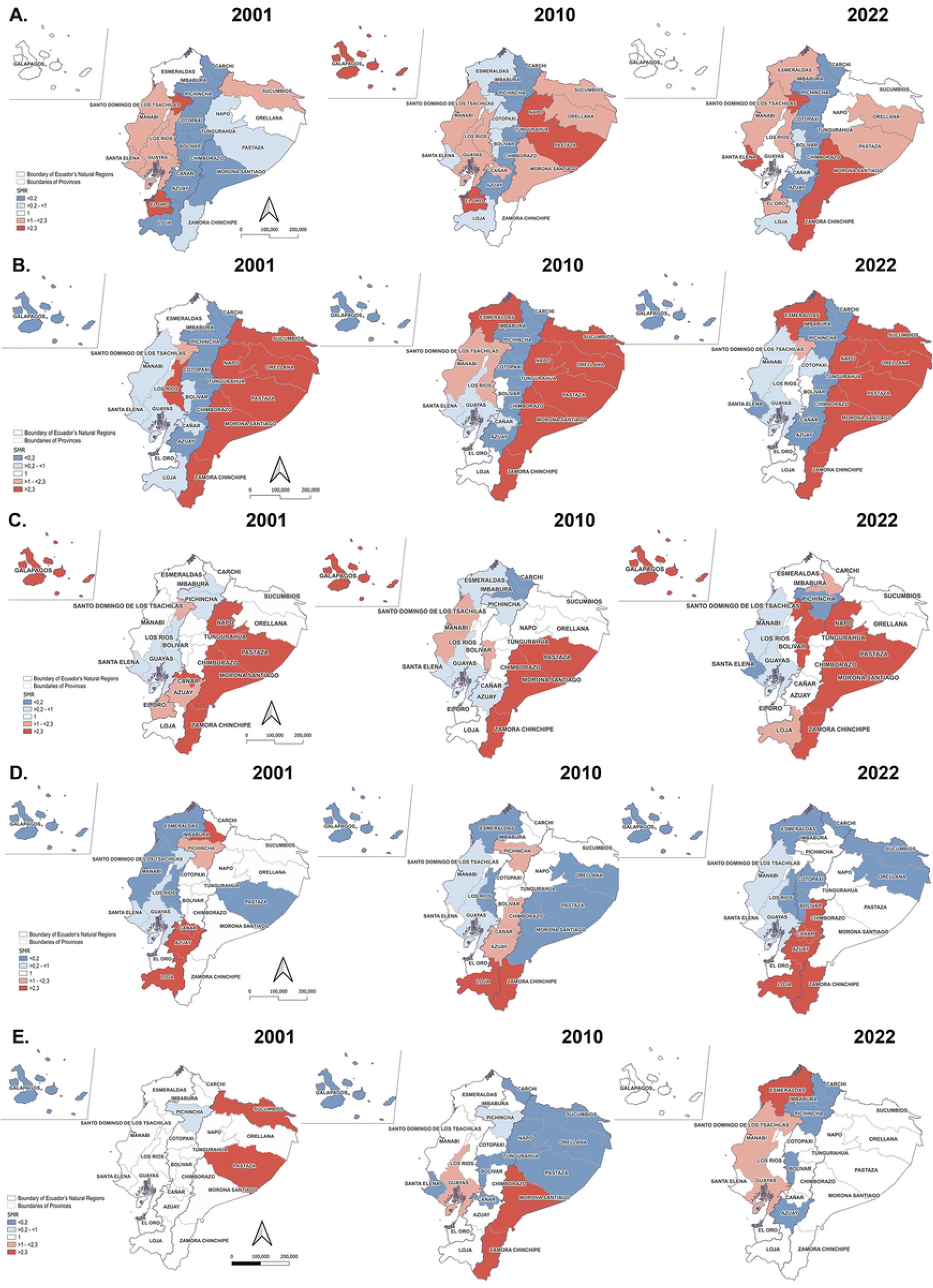
Standardised morbidity ratio (SMR) of five most frequent NTDs in Ecuador by geoclimatic region and province in census years of 2001 (left), 2010 (centre), and 2022 (right). The SMR (Standardised morbidity ratio) calculated using the indirect method is shown, based on the population of the three census years. Dengue / chikungunya (A), Snakebite envenoming (B), Soil-transmitted helminths (C), Taeniasis / cysticercosis (D), Scabies and other ectoparasitoses (E).

## DISCUSSION

This analysis of national hospitalization data offers the most comprehensive assessment to date of hospitalization trends for NTDs in Ecuador, capturing 25 years of data from 2000 to 2024. Our findings show an overall trend of increasing NTD hospitalizations explained by increasing arboviral morbidity (dengue and chikungunya), but otherwise significant reductions in non-arboviral NTD hospitalizations, the latter likely reflecting improvements in living conditions and health care access over this period.

The vast majority (97.1%) of NTD hospitalizations were attributable to five NTDs— arboviral infections (dengue/chik), snakebite envenoming, soil-transmitted helminthiases, taeniasis/cysticercosis, and ectoparasitoses— and reflects broader regional trends observed elsewhere in Latin America, including in northeastern Brazil, although with notable differences in disease burden composition. For example, while Brazil’s NTD hospitalization burden saw a decline of 10.3% over the period, 2001 to 2018, it prominently included leishmaniasis (8.6%) and leprosy (6.4%) [37], diseases that accounted for <1% of Ecuador’s NTD-related hospitalizations. Hospitalization rates for leprosy in Ecuador have declined from a rate of 1.33 (95% CI 1.06-1.60) per 100,000 in 2000 to 0.06 (0.01-0.11) in 2024 reflecting a declining disease incidence [38], while the cutaneous form of leishmaniasis that is endemic in Ecuador rarely requires hospitalization [39].

Arboviral infections (dengue/chik) represented the largest and fastest-growing share of hospitalizations, with an AAPC that increased steeply after 2018 (+47.8%). This rise reflects regional dengue trends [40]. The decline in dengue/chik hospitalizations around 2017 reflected a decrease in case numbers observed elsewhere in Latin America that has been explained by several factors including increased population immunity from previous years of high transmission, potential cross-protection from the Zika virus, and enhanced vector control efforts [41,42]. Seasonality analysis confirmed well-documented transmission patterns, with peaks in the rainy season, in agreement with previous studies from Ecuador and other regions of Latin America [18,43].

While arboviral NTD morbidity is increasing, that attributed to other non-arboviral NTDs such as STH, taeniasis/cysticercosis, and snakebite showed significant and sustained declines. For example, STH-related hospitalization decreased with an AAPC of -5.62%, suggesting the impact of deworming and sanitation programs, though these reductions are not uniform across demographic groups. Severe STH morbidity resulting in hospitalization results from complications of heavy parasite burdens such as intestinal obstruction in children caused by *Ascaris lumbricoides*, a leading cause of surgical emergencies [44], or severe anaemia consequent to hookworm infections [45]. The effects of deworming programmes and improving access to clean water and sanitation has been to reduce worm burdens such that increasingly small proportions of those infected are at risk of severe morbidity [16,46]. Women had higher hospitalization rates for STH than men, particularly from age 10 onward, consistent with prior studies indicating maternal and early childhood STH transmission risks [47,48].

Ectoparasitoses, particularly scabies, emerged as a growing cause of hospitalization, especially among the elderly. This aligns with trends seen in high-income countries such as Spain and South Korea [49,50]. The increase highlights the need for attention to geriatric NTD care and associated socio-environmental risk factors such as overcrowding, poverty, and poor hygiene.

Gender and age disparities were notable across NTDs. Snakebite disproportionately affected young males, as reported elsewhere across LMICs [51], likely due to occupational exposure in agriculture and forest-related work. Conversely, the rise in hospitalizations among older adults—especially from STH and ectoparasitoses— emphasizes the need for age-sensitive interventions and surveillance.

Geographically, the Amazon region consistently reported the highest SMRs for multiple NTDs, particularly snakebite, STH, and arboviruses. These provinces also exhibit the highest poverty rates, rurality, and proportions of Indigenous populations [25], exacerbating vulnerabilities to NTDs. The shift in arboviral SMR from the coastal region in earlier years to the Amazon by 2022 underscores how urbanization without adequate infrastructure (e.g., water, sanitation, and waste systems) facilitates vector proliferation [21]. Moreover, endemic persistence of taeniasis/cysticercosis in southern Andean provinces—associated with traditional pork consumption and limited meat inspection— confirms long-standing epidemiological patterns [22].

Despite Ecuador’s reported progress toward elimination of certain NTDs like yaws [15], and onchocerciasis whose elimination was certified in 2015 [13], our data show hospitalizations still being recorded for these and other “non-endemic” NTDS including Buruli ulcer. The presence of hospitalizations from ‘eliminated’ or non-endemic NTDs also reflects possible imported cases, diagnostic overlap/and or confusion with similar conditions (e.g., cutaneous leishmaniasis and syphilitic ulcers). These data highlight weaknesses in national health information systems and the need for improved ICD-10 coding practices and data quality control, clinical training, and diagnostic protocols. In the case of schistosomiasis and lymphatic filariasis (S3 Table), although published reports using these official data sources are available [52,53], to our knowledge, no case of autochthonous transmission of either infection (i.e. *Schistosoma* spp. and *Wuchereria bancrofti*) has been documented (i.e., with appropriate identification of the causative pathogen) in the country. Some of these, if true, may represent imported cases from other regions of Latin America or elsewhere these diseases are known to occur. Ecuador is crossed by the land routes of a growing influx of migrants on their way to the North. However, almost all these cases were documented among individuals with a normal residence in Ecuador (S4 table) and there was no evidence of an increase in cases over time.

The study’s observed overall low mortality rate of 0.4% for NTD hospitalizations belies important disease-specific disparities. Chagas (6.7%) and leprosy (3.8%) accounted for the highest case-fatality rates, in line with findings from Brazil and other countries [54]. The anomalous mortality rate given for rabies (8.4%) likely represents the reason for hospital admission (e.g., patients with animal bites considered at risk of rabies) rather than diagnosis at discharge. An isolated outbreak of rabies (causing non-hospitalized deaths) transmitted through the bites of hematophagous bats was reported from Indigenous communities in the Amazon region in 2011 [55]. Ecuador’s in-hospital dengue case fatality rate of 0.3% exceeds the national target of <0.04% [56] by almost ten-fold but meets the WHO goal of 0.5% for 2025 [5], although inclusion of deaths during hospitalization only likely underestimates mortality.

Limitations of this study include reliance on hospitalization data alone, with no access to outpatient or surveillance datasets that are not yet publicly available, and lack of diagnostic validation or comorbidity capture in the INEC database. Further, there appear to be important issues with data quality control with no independent verification of ICD-10 code accuracy or diagnostic validity. Some NTDs require confirmation through laboratory or imaging testing - such capacity is restricted to the larger cities and is not readily available in district hospitals in many parts of the country. Additionally, demographic variables such as ethnicity and rurality have only been available since 2014–2015, restricting our ability to consider these factors. Available data were aggregated and the analysis ecological, thus limiting our ability to account for multiple hospitalizations of the same individuals or infer from group-level associations to those of individuals. Despite these limitations, the study offers important insights into spatiotemporal trends of the country’s NTD disease burden that are relevant to informing public health policies and resource allocation. While, our data provides useful insights into spatiotemporal trends of the more severe spectrum of disease caused by NTDs (i.e. requiring hospitalization), the documentation of progress towards SDG 3.3 (i.e., case numbers) will require the national surveillance database to be made available to the academic community for analysis.

In conclusion, our data reveal increasing trends over time in hospitalizations attributed to arboviral infections and ectoparasites, in parallel, with declining hospitalization rates attributable to helminthiases and zoonotic parasitic infections. The latter likely reflect progressive improvements in health and sanitary infrastructure, and in economic and living conditions in the country since 2000. Comparisons in hospitalization rates between provinces and by age showed a relatively greater burden of more severe disease attributable to NTDs in the Amazon region and among vulnerable age groups. Documenting progress towards SDG 3.3 will require the national surveillance and primary care databases to be made available to the academic community. Interventions to control and eliminate NTDs likely will need to be geographically and demographically tailored—particularly to Amazonian populations and vulnerable age groups—and supported by stronger surveillance and more robust notification processes with improved diagnostic capacity, improved data collection and registration practices, and sustained health system investment.

## Data Availability

All data used in this study are available as a supporting information file.

## Author contributions

Conceptualization: PJC, CA, NRS. Data Curation: CA, PA, DZ. Formal Analysis: CA, PA, DZ, NRS, PJC. Funding Acquisition: PJC, NRS. Methodology: CA, PA, DZ, MM. Project Administration: NRS, PJC. Resources: NRS, PJC. Software: PA, DZ, GD. Supervision: PJC, NRS, MM. Visualization: PA, GD. Writing: CA, NRS, PJC. Writing – Review & Editing: All authors.

## Supporting information

**S1_Table. ICD-10 codes used for the 21 neglected tropical diseases**

**S2_Table. Age-standardized hospitalization rates for neglected tropical diseases (NTDs) in Ecuador between 2000 and 2024.** Shown are rates per 100,000 population and 95% confidence intervals.

**S3_Table. Frequency of hospital discharges for 7 non-endemic NTDs with corresponding hospitalizations reported in Ecuador between 2000 and 2024.**

**S4_Table. Frequencies and origins (area of usual residence) of patients hospitalized with non-endemic NTDs in Ecuador between 2015 and 2024 for which data are available.**

**S5_Table. Standardized morbidity ratio (and 95% confidence intervals) of hospitalization rates attributed to 5 most frequent endemic neglected tropical diseases in Ecuador by geoclimatic region and province in census years of 2001, 2010, and 2022.**

**S1_Fig. Age-standardized hospitalization rates for 14 endemics neglected tropical diseases (NTDs) in Ecuador between 2000 and 2024 (data are from S2_Table).**

**S2_Fig. Crude cumulative hospitalization rates (per 100,000 population) for endemic NTDs by province in Ecuador between 2000 and 2024.**

**S1 Data. Raw data used for analyses**

## References

1. World Health Organization. Global report on neglected tropical diseases 2023. 2023. Available: https://iris.who.int/bitstream/handle/10665/365729/9789240067295-eng.pdf?sequence=1

2. Ogieuhi IJ, Ajekiigbe VO, Aremu SO, Okpujie V, Bassey PU, Babalola AE, et al. Global partnerships in combating tropical diseases: assessing the impact of a U.S. withdrawal from the WHO. Trop Med Health. 2025;53: 36. doi:10.1186/s41182-025-00722-8

3. World Health Organization. Control of Neglected Tropical Diseases. 2017 [cited 19 Feb 2025]. Available: https://www.who.int/teams/control-of-neglected-tropical-diseases

4. Mahapatra B, Mukherjee N, Khatoon S, Bhattacharya P, Das P, Bharti O, et al. Economic evaluations of neglected tropical disease interventions in low- and middle-income countries: a systematic review protocol. JBI Evid Synth. 2024;22: 1582–1593. doi:10.11124/JBIES-23-00339

5. World Health Organization. Ending the neglect to attain the sustainable development goals: a sustainability framework for action against neglected tropical diseases 2021-2030. Geneva: World Health Organization; 2021. Available: https://iris.who.int/handle/10665/338886

6. World Health Organization. World health statistics 2024. 21 May 2025 [cited 30 Apr 2025]. Available: https://www.who.int/data/gho/whs-annex/

7. Liblik K, Byun J, Saldarriaga C, Perez GE, Lopez-Santi R, Wyss FQ, et al. Snakebite Envenomation and Heart: Systematic Review. Curr Probl Cardiol. 2021;47: 100861. doi:10.1016/j.cpcardiol.2021.100861

8. Sharaf MS. Scabies: Immunopathogenesis and pathological changes. Parasitol Res. 2024;123: 149. doi:10.1007/s00436-024-08173-6

9. Burgos LM, Farina J, Liendro MC, Saldarriaga C, Liprandi AS, Wyss F, et al. Neglected Tropical Diseases and Other Infectious Diseases Affecting the Heart. The NET-Heart Project: Rationale and Design. Glob Heart. 2020;15. doi:10.5334/gh.867

10. Araiza-Garaygordobil D, García-Martínez CE, Burgos LM, Saldarriaga C, Liblik K, Mendoza I, et al. Dengue and the heart. Cardiovasc J Afr. 2021;32: 46–53. doi:10.5830/CVJA-2021-033

11. Assis TM de, Rabello A, Cota G. Economic evaluations addressing diagnosis and treatment strategies for neglected tropical diseases: an overview. Rev Inst Med Trop Sao Paulo. 2021;63. doi:10.1590/s1678-9946202163041

12. Ministerio de Salud Pública. Manual de procedimientos del Subsistema de Vigilancia Epidemiológica alerta acción SIVE - ALERTA. Quito; 2014. Available: https://aplicaciones.msp.gob.ec/salud/archivosdigitales/documentosDirecciones/dnn/archivos/MANUAL%20DE%20PROCEDIMIENTOS%2016%20de%20Octubre%20de%202014.pdf

13. Guevara Á, Lovato R, Proaño R, Rodriguez-Perez MA, Unnasch T, Cooper PJ, et al. Elimination of onchocerciasis in Ecuador: findings of post-treatment surveillance. Parasit Vectors. 2018;11: 265. doi:10.1186/s13071-018-2851-3

14. Anselmi M, Moreira J, Caicedo C, Guderian R, Tognoni G. Community participation eliminates yaws in Ecuador. Tropical Medicine & International Health. 2003;8: 634–638. doi:10.1046/j.1365-3156.2003.01073.x

15. Cooper PJ, Anselmi M, Caicedo C, Lopez A, Vicuña Y, Cagua Ordoñez J, et al. Yaws elimination in Ecuador: Findings of a serological survey of children in Esmeraldas province to evaluate interruption of transmission. PLoS Negl Trop Dis. 2022;16: e0010173. doi:10.1371/journal.pntd.0010173

16. Moncayo AL, Lovato R, Cooper PJ. Soil-transmitted helminth infections and nutritional status in Ecuador: findings from a national survey and implications for control strategies. BMJ Open. 2018;8: e021319. doi:10.1136/bmjopen-2017-021319

17. Calvopina M, Ortiz-Prado E, Castañeda B, Cueva I, Rodriguez-Hidalgo R, Cooper PJ. Human myiasis in Ecuador. PLoS Negl Trop Dis. 2020;14: e0007858. doi:10.1371/journal.pntd.0007858

18. Sippy R, Herrera D, Gaus D, Gangnon RE, Patz JA, Osorio JE. Seasonal patterns of dengue fever in rural Ecuador: 2009-2016. PLoS Negl Trop Dis. 2019;13: e0007360. doi:10.1371/JOURNAL.PNTD.0007360

19. Lapo-Talledo GJ. Dengue hospitalizations and in-hospital mortality changes in trend in Ecuador: a nationwide study from 2015 to 2022. Infect Dis. 2024;56: 632–643. doi:10.1080/23744235.2024.2341871

20. Acosta-España JD, Dueñas-Espín I, Grijalva Narvaez DF, Altamirano-Jara JB, Gómez-Jaramillo AM, Rodriguez-Morales AJ. Analysis of inpatient data on dengue fever, malaria and leishmaniasis in Ecuador: A cross-sectional national study, 2015–2022. New Microbes New Infect. 2024;60–61: 101421. doi:10.1016/J.NMNI.2024.101421

21. Katzelnick LC, Quentin E, Colston S, Ha TA, Andrade P, Eisenberg JNS, et al. Increasing transmission of dengue virus across ecologically diverse regions of Ecuador and associated risk factors. PLoS Negl Trop Dis. 2024;18: e0011408. doi:10.1371/JOURNAL.PNTD.0011408

22. Rodriguez-Hidalgo R, Benitez-Ortiz W, Praet N, Saa L, Vercruysse J, Brandt J, et al. Taeniasis-cysticercosis in Southern Ecuador: assessment of infection status using multiple laboratory diagnostic tools. Mem Inst Oswaldo Cruz. 2006;101: 779–782. doi:10.1590/S0074-02762006000700012

23. Núñez-González S, Gault C, Simancas-Racines D. Spatial analysis of dengue, cysticercosis and Chagas disease mortality in Ecuador, 2011-2016. Trans R Soc Trop Med Hyg. 2019;113: 44–47. doi:10.1093/trstmh/try106

24. Toalombo Espin CJ, Coque Procel M. Leishmaniasis en el Ecuador: revisión bibliográfica. Mediciencias UTA. 2021;5: 2–11. doi:10.31243/mdc.uta.v5i3.1190.2021

25. Instituto Nacional de Estadísticas y Censos. Resultados principales del Censo Ecuador 2022. 2023 [cited 23 Feb 2025]. Available: https://censoecuador.ecudatanalytics.com/

26. Ministerio de Ambiente del Ecuador. Sistema de clasificación de los Ecosistemas del Ecuador Continental. Quito; 2012. Available: https://www.ambiente.gob.ec/wp-content/uploads/downloads/2012/09/LEYENDA-ECOSISTEMAS_ECUADOR_2.pdf

27. Instituto Nacional de Estadística y Censos. Boletín Registro Estadístico de Camas y Egresos Hospitalarios Año 2022. 2023. Available: https://www.ecuadorencifras.gob.ec/documentos/web-inec/Estadisticas_Sociales/Camas_Egresos_Hospitalarios/Cam_Egre_Hos_2022/ Presentacion_ECEH_2022.pdf

28. World Health Organization. Neglected tropical diseases. 2025 [cited 17 Jul 2025]. Available: https://www.who.int/health-topics/neglected-tropical-diseases#tab=tab_1

29. Instituto Nacional de Estadística y Censos. Registro estadístico de Camas y Egresos Hospitalarios. 2025 [cited 2 Sep 2025]. Available: https://www.ecuadorencifras.gob.ec/camas-y-egresos-hospitalarios/

30. World Health Organization. Official WHO ICD-10 Updates 2019 Package. 1 Feb 2020 [cited 18 May 2025]. Available: https://www.who.int/publications/m/item/official-who-icd-10-updates-2019-package

31. Ahmad O Ben, Boschi Pinto C, Lopez AD. Age Standardization of Rates: A New WHO Standard. GPE Discussion Paper Series: No 31. 2001; 10–12. Available: https://cdn.who.int/media/docs/default-source/gho-documents/global-health-estimates/gpe_discussion_paper_series_paper31_2001_age_standardization_rates.pdf

32. Kim HJ, Fay MP, Feuer EJ, Midthune DN. Permutation tests for joinpoint regression with applications to cancer rates. Stat Med. 2000;19: 335–351. doi:10.1002/(SICI)1097-0258(20000215)19:3<335::AID-SIM336>3.0.CO;2-Z

33. Lal A, Hales S, French N, Baker MG. Seasonality in Human Zoonotic Enteric Diseases: A Systematic Review. PLoS One. 2012;7: e31883. doi:10.1371/journal.pone.0031883

34. Zhao D, Zhang H, Wu X, Zhang L, Li S, He S. Spatial and temporal analysis and forecasting of TB reported incidence in western China. BMC Public Health. 2024;24: 2504. doi:10.1186/s12889-024-19994-6

35. Instituto Nacional de Estadística y Censos. Estimaciones y Proyecciones de Población. 2024 [cited 12 Sep 2024]. Available: https://www.ecuadorencifras.gob.ec/proyecciones-poblacionales/

36. Instituto Nacional de Estadística y Censos. Población y Demografía. [cited 26 May 2025]. Available: https://www.ecuadorencifras.gob.ec/censo-de-poblacion-y-vivienda/

37. Brito SP de S, Lima M da S, Ferreira AF, Ramos Jr AN. Hospital admissions due to neglected tropical diseases in Piauí, in the Northeast region of Brazil: costs, time trends, and spatial patterns, 2001-2018. Cad Saude Publica. 2022;38: e00281021. doi:10.1590/0102-311XPT281021

38. Hernandez-Bojorge S, Gardellini T, Parikh J, Rupani N, Jacob B, Hoare I, et al. Ecuador Towards Zero Leprosy: A Twenty-Three-Year Retrospective Epidemiologic and Spatiotemporal Analysis of Leprosy in Ecuador. Trop Med Infect Dis. 2024;9: 246. doi:10.3390/tropicalmed9100246

39. Hashiguchi Y, Velez LN, Villegas N V., Mimori T, Gomez EAL, Kato H. Leishmaniases in Ecuador: Comprehensive review and current status. Acta Trop. 2017;166: 299–315. doi:10.1016/j.actatropica.2016.11.039

40. Lin Y, Fang K, Zheng Y, Wang H-L, Wu J. Global burden and trends of neglected tropical diseases from 1990 to 2019. J Travel Med. 2022;29. doi:10.1093/jtm/taac031

41. Perez F, Llau A, Gutierrez G, Bezerra H, Coelho G, Ault S, et al. The decline of dengue in the Americas in 2017: discussion of multiple hypotheses. Tropical Medicine & International Health. 2019;24: 442–453. doi:10.1111/tmi.13200

42. Liang Y, Dai X. The global incidence and trends of three common flavivirus infections (Dengue, yellow fever, and Zika) from 2011 to 2021. Front Microbiol. 2024;15. doi:10.3389/fmicb.2024.1458166

43. Borges IVG, Musah A, Dutra LMM, Tunali M, Lima CL, Tunali MM, et al. Analysis of the interrelationship between precipitation and confirmed dengue cases in the city of Recife (Brazil) covering climate and public health information. Front Public Health. 2024;12. doi:10.3389/fpubh.2024.1456043

44. Else KJ, Keiser J, Holland C V, Grencis RK, Sattelle DB, Fujiwara RT, et al. Whipworm and roundworm infections. Nat Rev Dis Primers. 2020;6: 44. doi:10.1038/s41572-020-0171-3

45. Loukas A, Hotez PJ, Diemert D, Yazdanbakhsh M, McCarthy JS, Correa-Oliveira R, et al. Hookworm infection. Nat Rev Dis Primers. 2016;2: 16088. doi:10.1038/nrdp.2016.88

46. Chis Ster I, Niaz HF, Chico ME, Oviedo Y, Vaca M, Cooper PJ. The epidemiology of soil-transmitted helminth infections in children up to 8 years of age: Findings from an Ecuadorian birth cohort. PLoS Negl Trop Dis. 2021;15: e0009972. doi:10.1371/journal.pntd.0009972

47. Menzies SK, Rodriguez A, Chico M, Sandoval C, Broncano N, Guadalupe I, et al. Risk Factors for Soil-Transmitted Helminth Infections during the First 3 Years of Life in the Tropics; Findings from a Birth Cohort. PLoS Negl Trop Dis. 2014;8: e2718. doi:10.1371/journal.pntd.0002718

48. Romero-Sandoval N, Ortiz-Rico C, Sánchez-Pérez HJ, Valdivieso D, Sandoval C, Pástor J, et al. Soil transmitted helminthiasis in indigenous groups. A community cross sectional study in the Amazonian southern border region of Ecuador. BMJ Open. 2017;7: e013626. doi:10.1136/bmjopen-2016-013626

49. Redondo-Bravo L, Fernandez-Martinez B, Gómez-Barroso D, Gherasim A, García-Gómez M, Benito A, et al. Scabies in Spain? A comprehensive epidemiological picture. PLoS One. 2021;16: e0258780. doi:10.1371/journal.pone.0258780

50. Kim JH, Cheong HK. Epidemiologic Trends and Seasonality of Scabies in South Korea, 2010-2017. Korean J Parasitol. 2019;57: 399–404. doi:10.3347/KJP.2019.57.4.399

51. Afroz A, Siddiquea BN, Chowdhury HA, Jackson TN, Watt AD. Snakebite envenoming: A systematic review and meta-analysis of global morbidity and mortality. PLoS Negl Trop Dis. 2024;18: e0012080. doi:10.1371/journal.pntd.0012080

52. Vásconez-González J, Yeager J, Izquierdo-Condoy JS, Fernandez-Naranjo R, López M-B, Dávila MG, et al. An 11-year epidemiological analysis of schistosomiasis in Ecuador: Investigating a non-endemic, neglected, and challenging-to-identify parasitic disease. Food Waterborne Parasitol. 2023;31: e00196. doi:10.1016/j.fawpar.2023.e00196

53. Izquierdo-Condoy JS, Naranjo-Lara P, Vásconez-Gonzalez J, Fernandez-Naranjo R, Placencia-André R, Davila MG, et al. A nationwide epidemiological and geodemographic analysis of lymphatic filariasis in Ecuador: a neglected and often forgotten disease in Ecuador. Front Public Health. 2023;11: 1270015. doi:10.3389/fpubh.2023.1270015

54. Cucunubá ZM, Okuwoga O, Basáñez M-G, Nouvellet P. Increased mortality attributed to Chagas disease: a systematic review and meta-analysis. Parasit Vectors. 2016;9: 42. doi:10.1186/s13071-016-1315-x

55. Romero-Sandoval N, Parra C, Gallegos G, Guanopatín A, Campaña MF, Haro M, et al. Haematophagous bat bites in Ecuadorian Amazon: characterisation and implications for sylvatic rabies prevention. Public Health Action. 2013;3: 85–89. doi:10.5588/pha.12.0070

56. Ministerio de Salud Pública. Plan Decenal de Salud 2022 2023. MSP. 2022.

